# Electrophysiological Markers of Within-Network Connectivity in Major Depression

**DOI:** 10.64898/2026.08.04.26359708

**Authors:** Yoojin Lee, Elizabeth D. Ballard, Jeffrey D. Stout, Allison Nugent, Hiroe Hu, Kelly T. Hurst, Amy Xu, Carlos A. Zarate, Jessica R. Gilbert

## Abstract

Depression and treatment-resistant depression (TRD) are significant public health issues, but the associated network-level neurobiological mechanisms remain poorly understood. This study used magnetoencephalography (MEG) to identify altered resting-state connectivity within the default mode (DMN), executive control (ECN), salience (SN), dorsal attention (DAN), motor (MN), and visual (VN) networks as potential biomarkers of depression and treatment resistance. The study recruited 168 participants (80 healthy volunteers (HVs) and 88 currently experiencing a major depressive episode (74 with TRD and 14 without TRD (noTRD))). Data Integration Analysis for Biomarker Discovery using Latent Variable Approaches for Omics Studies (DIABLO) was used to differentiate the depression, TRD, and HV subgroups and identify neural markers of depression and treatment resistance. For differentiating the depression and HV groups, the triple network model (area under the receiver operating curve (AUROC): 0.759- 0.787)—which includes the DMN, ECN, and SN—outperformed the six-network model (AUROC: 0.747-0.762) across different bandwidths. For differentiating the TRD and HV groups, the triple network model demonstrated reasonable prediction across different bandwidths (AUROC: 0.737-0.807); potential within-network connectivity differences distinguished those with TRD from HVs, especially DMN within-network connectivity between the inferior parietal lobule and precuneus in the beta band (*FDR-corrected p*<.05). Hyperconnectivity within the SN (superior parietal lobule and frontal operculum in the alpha band) and DMN (inferior parietal lobule and lateral prefrontal cortex in the beta band) was associated with number of treatment failures (ps<.05). These findings highlight key brain regions and connectivity patterns, advancing our understanding of neural mechanisms underlying depression and treatment resistance.

## Introduction

Of the more than 280 million people worldwide who suffer from depression [1], 30% do not respond to standard pharmacological treatments and develop treatment-resistant depression (TRD), which imposes significant burdens [2]. Although diagnosis and monitoring are still largely based on clinician-administered questionnaires, heterogeneity and the complexity of depression, particularly TRD, highlight the need for objective neurobiological markers. In this context, emerging evidence suggests that altered brain network connectivity occurs in depression and TRD [3, 4]. Advancing our understanding of these disorders through the lens of brain network connectivity holds promise for improving diagnostic precision and informing the development of targeted interventions.

Distinct brain connectivity patterns have been identified in depression and TRD across canonical networks, including the default mode network (DMN), the executive control network (ECN), and the salience network (SN) (i.e., the “triple network” model) [5, 6]. However, evidence for depression-associated changes in the DMN specifically remains mixed, particularly in TRD patients. For instance, Guo and colleagues [7] found reduced intra-DMN connectivity in both individuals with depression and those with TRD, but other studies of patients with major depressive disorder (MDD) found the opposite result [8–11]. Furthermore, in TRD patients specifically, some studies reported increased intra-DMN connectivity, while others observed diminished functional connectivity both within the DMN and with other networks [12–15].

Taking heterogeneity across studies into account, a systematic review also concluded that TRD is most consistently associated with reduced DMN functional connectivity [16]. Notably, a recent study revealed connectivity between two DMN hubs—the rostral anterior cingulate cortex and the posterior cingulate cortex—in individuals with TRD compared to those with treatment- responsive depression and healthy volunteers (HVs) [13]. Beyond the DMN, both depression and TRD have been associated with ECN hypoconnectivity—especially between the dorsolateral prefrontal cortex (DLPFC) and the angular gyrus or inferior parietal lobule (IPL) [12, 14], although ECN hyperconnectivity has also been observed [17]. With regard to the SN, studies identified reduced connectivity in individuals with depression [14, 18–20], including TRD [14].

In addition to the triple network model, a growing body of research suggests that brain connectivity within other networks—including the dorsal attention network (DAN), motor network (MN), and visual network (VN)—may also be dysregulated in depression, including TRD. In depression, connectivity within the DAN appeared less internally coherent, with reduced network homogeneity evident in the supramarginal gyrus [21, 22]. Altered connectivity in the MN and VN, especially associated with movement and visual processing, has also been linked to psychomotor retardation in TRD patients and may serve as a marker of depression [23–25].

Taken together, these studies indicate that network-level alterations are consistently implicated in depression, particularly TRD. However, the exact patterns and inter-relationships remain unanswered, underscoring the need for more integrative network-level investigations. Notably, most findings on connectivity in these patient populations come from functional magnetic resonance imaging (fMRI) studies with relatively few TRD patients that measured hemodynamic signals over seconds. This limited temporal resolution obscures rapid neural connections, leaving the time-resolved dynamics of these networks largely unexplored. Because connections within a given network are more likely to share functional properties than connections between networks, one potential approach is to examine within-network connectivity, which may capture functionally specific aspects of depression, particularly TRD.

At the same time, averaging connectivity across entire networks may obscure meaningful regional heterogeneity, thereby complicating clinical interpretation. These considerations highlight the need to investigate within-network connectivity across the networks implicated in depression and TRD while accounting for both temporal dynamics and regional heterogeneity within each network.

Electrophysiological techniques such as electroencephalography (EEG) and magnetoencephalography (MEG) offer high-resolution temporal mapping of neural electrical currents with millisecond precision. Although electrophysiological modalities have been used to study depression, they remain relatively underutilized in investigating TRD. A handful of studies that investigated resting-state EEG power differences found that depression was associated with changes in low-frequency power. For example, elevated theta, alpha, and beta power was observed in both anterior [26, 27] and posterior brain regions [28] in treatment-responsive MDD patients. Within-network connectivity has also been examined in the context of studying canonical network dysfunction in depression and response to treatment. Reduced MEG-derived subgenual anterior cingulate to hippocampal connectivity coupled with increased insulo-temporal to amygdala connectivity in the beta frequency was observed in MDD patients compared to HVs [29]. In terms of treatment response, individuals with remitted depression were found to exhibit increased EEG-measured beta within-network connectivity in the DMN, ECN, and SN compared to both HVs and those with non-remitted MDD [30]. Collectively, these findings suggest that dysregulated signals, particularly in the theta, alpha, and beta bands, could help distinguish individuals with depression, including those with TRD, from HVs.

The present study addresses previous research gaps by examining MEG-derived within-network connectivity across specific frequency bands in a comparatively large sample of individuals with TRD. Data Integration Analysis for Biomarker discovery using Latent variable approaches for Omics studies (DIABLO) was used to determine whether canonical brain networks could differentiate individuals with depression from those with TRD and from HVs (Fig. 1). Whereas prior neuroimaging studies examining connectivity in TRD have typically been constrained by small sample sizes (range: N=17-38) [7, 12–14, 20], our larger cohort (n=88), all of whom were recruited and studied under consistent clinical conditions and procedures, allows investigation of neural features associated with specific symptom dimensions rather than depression more broadly. DIABLO facilitates the identification of a focused set of correlated predictors spanning canonical networks and frequency bands, thus enabling evaluation of whether within-network connectivity (estimated using publicly available atlases [31]) could differentiate individuals with depression and TRD and whether these group differences varied by symptom severity. Canonical networks of interest included the DMN, ECN, SN, DAN, MN, and VN. The first question of interest was whether group differences in depression (compared to HVs) could be better predicted using a six-network model or a three-network model (DMN, ECN, SN). The more robust network model was then applied to assess band-limited within-network connectivity features that differentiated individuals with depression or TRD from HVs. The key connectivity features underlying these group differences within each network were also investigated.

**Figure 1.**
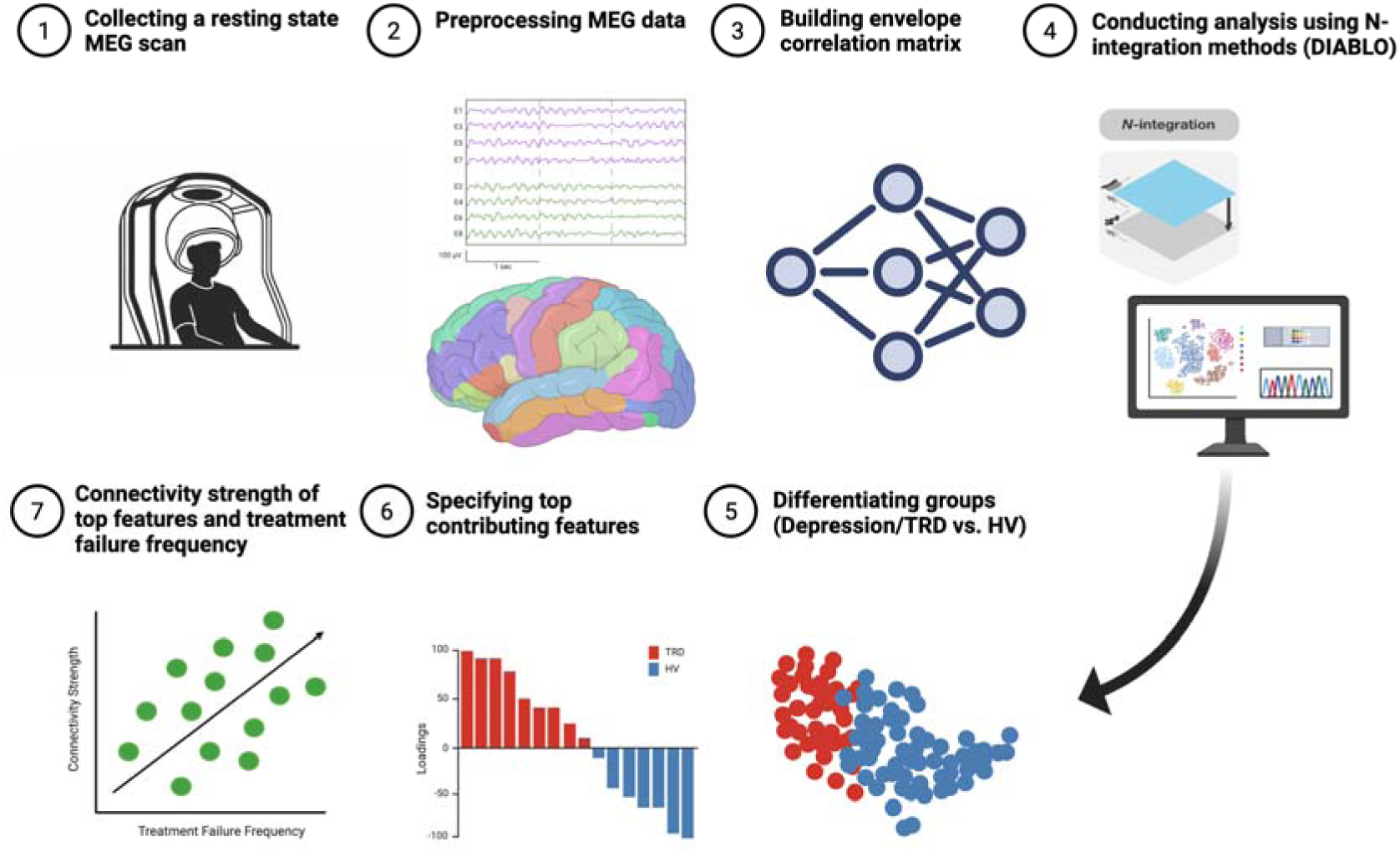
Study Design. The current study included the following steps: (1) collection of resting-state magnetoencephalography (MEG) scans from participants; (2) preprocessing MEG data using the MNE Python pipeline; (3) building the brain connectivity matrix using the envelope correlation method; (4) conducting N-integration based sparse generalized canonical correlation analysis (DIABLO) to differentiate between the depression or treatment-resistant depression (TRD) group and the healthy volunteer (HV) group; (5-6) after specifying the groups, estimating which top features of connectivity strength contributed to the differentiation; and (7) estimating the connectivity strength of selected features and their relationship to the frequency of treatment failures.

## Materials and Methods

### Participants

Participants (n=181; 96 with depression and 85 HVs) were aged 19-65 years and had at least one resting-state MEG scan. All participants had enrolled in one of several clinical trials conducted at the National Institute of Mental Health in Bethesda, MD, USA (NCT00088699, NCT00369915, NCT02543983, NCT00397111, NCT03065335, NCT04821271, NCT00024635, NCT00472576, NCT00759395, NCT02049385, or clinical protocol 09-M-0240). Although participants were recruited across multiple cohorts, all data were collected under standardized protocols that used identical measurement instruments, controlled inpatient conditions, and uniform clinical characterization procedures. Detailed inclusion and exclusion criteria are provided in the Supplement.

Four participants (three with depression and one HV) were subsequently excluded due to poor imaging quality. Among the remaining 177 participants, an additional nine participants (two with TRD, three in the noTRD group, and four HVs) were excluded because of extreme envelope correlation scores across the networks; these were visually identified by extreme eigenvalues in the principal component analysis (PCA). The final dataset thus included 168 participants (n=88 with depression (87 with MDD and one with bipolar disorder) and 80 HVs.

Psychiatric diagnoses were confirmed using the Structured Clinical Interview for DSM Psychiatric Disorders (SCID) [32]. Depending on the treatment trial, participants could be either unmedicated or continuing to receive prescribed medications.

Participants with depression were further categorized into those who had TRD (n=74; 73 with MDD and one with bipolar disorder), defined as lack of response to two adequate trials of selective serotonin reuptake inhibitors (SSRIs), serotonin-norepinephrine reuptake inhibitors (SNRIs), or other antidepressants for the current major depressive episode, and those with depression but not TRD (noTRD, n=14 MDD patients). The predominance of TRD patients reflects that our protocols were designed primarily to investigate TRD, which is the primary focus of our research program. The noTRD group was nevertheless included because these individuals serve as a relevant comparison group. HVs had no personal or family history of psychiatric disorders as determined by the SCID. The mean age for the entire sample was 37.20 years (SD=12.50), and 48.81% were female (n=82). Demographic information appears in Table 1. The NIH Institutional Review Board approved the protocols, and all participants provided written, informed consent at the time of entry into their respective trials.

**Table 1.** Descriptive Statistics.

| Comparison Groups | Label | Sample Size | Age | Biological Sex (Female) | Body Mass Index | Ethnicity (White) | MADRS |
| --- | --- | --- | --- | --- | --- | --- | --- |
| **Depression vs HV** | Depression | 88 | 42.20 (13.00) | 41 (46.59) | 27.60 (7.79) | 71 (80.68) | 31.30 (8.40) |
|  | HV | 80 | 31.70 (9.32) | 41 (51.25) | 26.40 (4.55) | 45 (56.25) | 0.38 (0.75) |
| | Diff Stats | | $t(157.78) = -6.10, p < .001$ | $\chi^2 = 0.36, p = .645$ | $t(142.43) = -1.23, p = .220$ | $\chi^2 = 11.70, p = .002$ | $t(118.73) = -27.35, p < .001$ |
| | Mean Diff: Depression vs. HV | | 10.56, $p < .001$ | - | - | - | 30.92, $p < .001$ |
| **TRD vs noTRD vs HV** | TRD | 74 | 42.60 (13.00) | 36 (48.65) | 27.70 (8.37) | 62 (83.78) | 32.09 (7.73) |
|  | noTRD | 14 | 42.60 (13.01) | 5 (35.71) | 27.10 (3.56) | 9 (64.29) | 25.50 (10.52) |
|  | HV | 80 | 31.70 (9.32) | 41 (51.25) | 26.40 (4.55) | 45 (56.25) | 0.38 (0.75) |
| | Diff Stats | | $F(2,165) = 18.23, p < .001$ | $\chi^2 = 1.15, p = .570$ | $F(2,165) = 0.77, p = .466$ | $\chi^2 = 13.80, p = .001$ | $F(2,126) = 366.10, p < .001$ |
| | Mean Diff: TRD vs. HV | | 10.93, $p < .001$ | - | - | $\chi^2(1) = 12.50, p < .001$ | 31.71, $p < .001$ |
| | Mean Diff: TRD vs. noTRD | | 2.32, $p = .764$ | - | - | $\chi^2(1) = 1.76, p = .090$ | 6.59, $p = .007$ |
| | Mean Diff: noTRD vs. HV | | 8.60, $p = .027$ | - | - | $\chi^2(1) = 0.07, p = .575$ | 25.12, $p < .001$ |
| **TRD vs HV** | TRD | 74 | 42.60 (13.00) | 36 (48.65) | 27.70 (8.37) | 62 (83.78) | 32.10 (7.73) |
|  | HV | 80 | 31.70 (9.32) | 41 (51.25) | 26.40 (4.55) | 45 (56.25) | 0.38 (0.75) |
| | Diff Stats | | $t(131.39) = -5.95, p < .001$ | $\chi^2 = 0.10, p = .871$ | $t(110.72) = -1.18, p = .241$ | $\chi^2 = 13.74, p = .001$ | $t(69.78) = -33.85, p < .001$ |
| | Mean Diff: TRD vs. HV | | 10.93, $p < .001$ | - | - | - | 31.71, $p < .001$ |

TRD: treatment-resistant depression; noTRD: diagnosed with depression but not treatment-resistant depression; HV: healthy volunteer; MADRS:
Montgomery-Asberg Depression Rating Scale

### Neuroimaging acquisition: MEG

One or two eyes-closed resting-state MEG scans were collected during the baseline scanning session, which lasted eight minutes. Each scan lasted between 250 and 600 seconds, depending on the protocol. Neuromagnetic data were collected using a CTF 275-channel whole-head system using first-order axial gradiometer MEG sensors and superconducting quantum interference devices (VSM MedTech Ltd., Coquitlam, BC, Canada). Data were collected at 1200 Hz with a 0-300 Hz bandwidth, and synthetic third-order balancing was used to attenuate background environmental noise. In addition, T1-weighted magnetic resonance imaging scans were collected from each participant in a separate scanning session and co-registered to the MEG images using fiducial markers placed at the nasion, left, and right preauricular locations.

MEG data preprocessing was performed using the computational resources of the NIH Biowulf high-performance computing cluster (http://hpc.nih.gov).

### Resting-state image processing

Resting-state MEG data were processed using MNE python (ver 1.3 [33]). Source localization was performed using MNE python [33] along with utility functions in the nih_to_mne module (https://github.com/nih-megcore/nih_to_mne). MRI and MEG data were coregistered using the co-localized fiducial points, and boundary element method model solutions were also created using FreeSurfer [34, 35]. A linearly constrained minimum variance beamformer algorithm [36, 37] was used to obtain source localized data. LCMV beamforming is a spatial filtering technique that enhances signal-to-noise ratio by minimizing output variance while maintaining unit gain for a specific location, thus isolating desired signals and suppressing noise and interference. The estimated solution at each vertex was projected within multiple bandwidths, including the theta (4-8 Hz), alpha (9-14 Hz), beta (15-29 Hz), and gamma bands (30-58 Hz) [38]. The projected MEG power maps were then segmented into 200 parcels across 17 networks, as defined by Kong and colleagues [31]. These time series within each parcel were orthogonalized, and pairwise envelope correlation coefficients of those parcels were estimated using the Hilbert transform [39]. The correlation coefficients between parcels located within the same networks, including the DMN, ECN, SN, DAN, MN, and VN, were defined as within-network connectivities. For each network, amplitude envelope correlation values were mean-centered across within-network connectivity.

### DIABLO analysis

DIABLO (mixOmics, R v6.30.0) [40, 41] was used to analyze resting-state within-network brain connectivity metrics, defined as all possible pairwise connections between parcels within each network, across six networks, comprising 703 DMN, 780 ECN, 325 SN, 276 DAN, 276 MN, and 465 VN connectivity features. Supervised multi-block sparse partial least squares (sPLS) models with a tuned design matrix were applied to identify correlated variables distinguishing participant groups. Models were estimated separately for each frequency band because theta, alpha, beta, and gamma oscillations have been independently associated with distinct cognitive functions [42] and are known to arise from laminar-specific pathways [43] that mediate top-down (alpha/beta) and bottom-up signaling (gamma/theta) [44], which in turn contributes to network coherence [45]. Estimating within-network connectivity across multiple frequency bands simultaneously may conflate inhibitory and excitatory dynamics reflected in oscillatory MEG power. By analyzing each frequency band separately, our analysis preserved interpretability and avoided the risk of selecting only the strongest signal while ignoring frequency band-specific functional contributions, which could obscure frequency band-specific group differences. Detailed information is provided in the Supplemental Methods.

### Data Analysis Plan

To identify potential outliers, brain connectivity scores in each canonical network (DMN, ECN, SN, DAN, MN, and VN) were analyzed using PCA with the epPCA function in R [46].

The results were visualized with a scree plot, and individuals showing extreme eigenvalues along the principal components were excluded from further analysis. Within-network connectivity scores were standardized and entered as network-specific input blocks, and group membership was entered as the output in the DIABLO model.

To determine whether model performance was improved by emphasizing inter-network integration or class discrimination, two approaches for specifying the DIABLO design matrix were used to compare individuals with depression versus HVs using the triple-network model (DMN, ECN, and SN) across theta, alpha, beta, and gamma frequency bands. First, a sparse approach based on sPLS was used to model inter-network correlations while discriminating between groups. Second, a maximum discrimination approach was applied in which inter-network correlations were set to a minimal value (0.1) to maximize group discrimination. For each approach and frequency band, within-network connectivity scores from the DMN, ECN, and SN were entered as inputs and group membership (depression (which included both the TRD and noTRD groups) vs. HVs) as the outcome. The model was evaluated using the area under the receiver operating curve (AUROC) with centroid-based five-fold cross-validation repeated 100 times, implemented with the auroc function in the DIABLO package.

To determine whether group differentiation was adequately captured by the canonical triple network model (DMN, ECN, and SN) or benefited from more network coverage, the triple network model was compared with a six network model that additionally included the DAN, MN, and VN in different bandwidths (theta, alpha, beta, and gamma). This comparison tested whether discriminative information was centered within the triple networks canonically implicated in depression or distributed broadly across additional brain networks. These two models were compared across group contrasts (depression [TRD+noTRD] vs. HV; TRD vs noTRD vs HV; TRD vs HV) and different frequency bands (theta, alpha, beta, gamma). Model performance was evaluated using AUROC with the same cross-validation settings.

Because individuals with TRD comprised most of the depression group—potentially driving observed differences between the depression and HV groups—an additional exploratory analysis was conducted to better distinguish group-specific effects. Group membership was entered to differentiate between groups across different bandwidths (theta, alpha, beta, gamma): 1) TRD participants from HVs, and 2) TRD participants versus noTRD participants versus HVs using only the discrimination approach to maximize separation between groups. To compare predictive accuracy across networks within a given frequency band, an exploratory bootstrap approach with simulated receiver operating curve (ROC) values was used (see Supplemental Methods). This is because AUROC values were derived from DIABLO models that account for latent multivariate relationships among within-network connectivity features—relationships that cannot be directly reproduced from raw data. This approach accounts for sampling variability and provides a robust test of differences in predictive performance between the networks.

Multivariate analyses of variance (MANOVA) tests were conducted to verify the extent to which each brain connectivity score could differentiate group membership. Group membership was entered as an independent variable, and each brain connectivity score was entered as a dependent variable. Biological sex, age, and race/ethnicity were entered as covariates. To compare the predictability of each network, simulations were used to compare the AUROC scores of the two networks (for details, see Supplemental Methods). To account for multiple testing, false discovery rate (FDR) correction was applied to the omnibus group-effect p-values from the ANOVA models. Specifically, for each network-component analysis, p-values were corrected across the set of selected connectivity. FDR adjustment was performed using the Benjamini-Hochberg procedure. Corrected p-values are reported as FDR-adjusted p-values, and effects were considered significant at p<.05. Tukey’s Honestly Significant Difference (HSD) was used to adjust for multiple pairwise group comparisons within each connectivity feature.

Because the omnibus group effect was tested across multiple selected connectivity features, the resulting ANOVA p-values were additionally adjusted across features using the Benjamini-Hochberg FDR procedure.

An exploratory analysis examined whether selected features from the TRD model were quantitatively related to the frequency of treatment resistance. An ordered logistic regression analysis was conducted using the polr function from the MASS package in R [47]. Selected within-network connectivity features were entered as predictive variables, and quantile scores of treatment failure frequency were entered as outcome variables. Biological sex and age were included as covariates. Sensitivity analyses were conducted to determine whether the association was suppressed after adjusting for severity of depressive symptoms (assessed by Montgomery-Asberg Depression Rating Scale (MADRS) score), medication history (coded as 0 for any prescribed medication and 1 for no medication), or polypharmacy (number of prescribed medications).

## Results

### Descriptive statistics

The noTRD/TRD and HV groups did not differ significantly with regard to biological sex or body mass index (ps>.05; Table 1). Post-hoc analyses found that the depression, TRD, and noTRD groups were all older than the HV group (p<.05). Within the TRD group, 12 patients (16.20%) were prescribed medication during the trial. The frequency of polypharmacy across the group was low (M=0.45, SD=1.11).

### DIABLO model performance: sparse vs. discrimination and network comparisons

DIABLO models were first evaluated using a sparse approach that models correlations among networks while accounting for differences between them. These models demonstrated high inter-network correlations, which were subsequently entered as “weights” in the design matrix. Under this approach, the three-network model (DMN, ECN, SN) yielded modest classification performance (AUROC: 0.611 to 0.646 across bandwidths; Table S2).

In contrast, a maximum discrimination approach (design matrix = 0.1 [48]), reflecting minimal inter-network correlation, substantially improved performance (AUROC: 0.758-0.787 under the same cross-validation settings; Table S2). This approach was therefore used for additional exploratory sub-group analyses.

Applying the maximum discrimination framework, a six-network model (DMN, ECN, SN, DAN, MN, and VN) produced similar or slightly lower performance (AUROC: 0.747– 0.762, Table S1) than the three-network model. These findings suggest that discrimination between the depression and HV groups was primarily driven by the canonical triple network (DMN, ECN, SN), which was retained for all group-level analyses.

In addition, inter-component correlation, sample, and arrow plots indicated modest heterogeneity in group discrimination across networks (Fig .2a-c).

**Figure 2.**
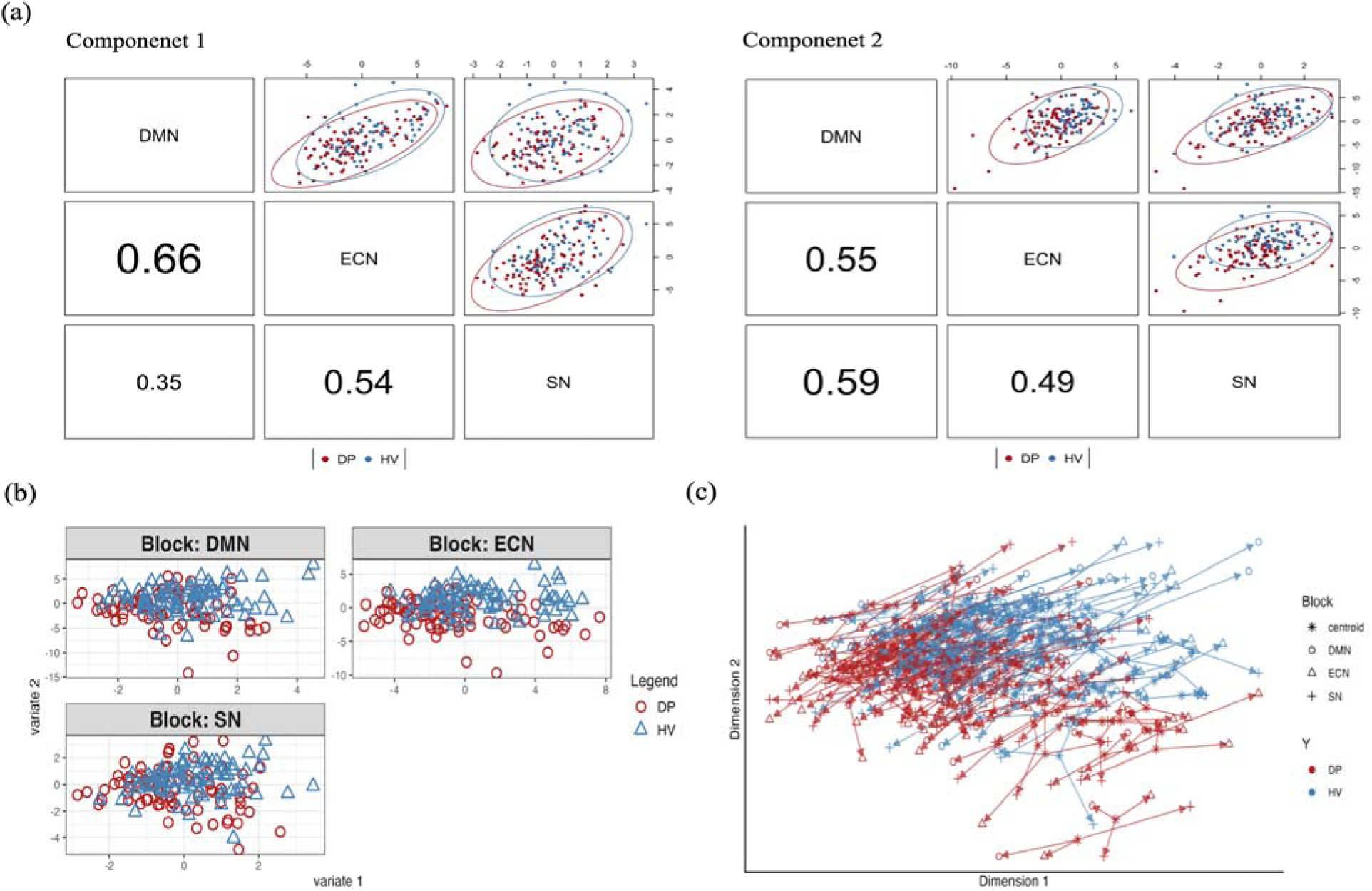
DIABLO results differentiating individuals with depression (DP) from healthy volunteers (HVs) using beta-band within-network connectivity derived from magnetoencephalography (MEG): (a) inter-component correlation plots of components 1 and 2; (b) sample plot; (c) arrow plot. DMN: default mode network; ECN: executive control network; SN: salience network.

### DIABLO: Group differences

Most individuals in the depression group (n=74; 84.09%) had moderate to severe depressive symptoms (Table 1). Exploratory analyses with three canonical networks were performed to explore DIABLO’s ability to differentiate the TRD, noTRD, and HV groups. Using five-fold cross-validation, 100 repetitions, and two specified components, the maximum discrimination model significantly distinguished the TRD group from the others (ps<.05) except in the alpha band (AUROC=.649, p=.089; Table 2). The model also differentiated the noTRD group in the alpha (AUROC=.892, p=.010) and beta (AUROC=.647, p=.021) bands and distinguished the HV group from the others (ps<.05) except in the theta band (AUROC=.670, p=.072). To further clarify these findings, a DIABLO analysis was conducted that compared only the TRD and HV groups, which yielded AUROC scores ranging from 0.737 to 0.807, comparable to those from the depression versus HV comparison, which ranged from 0.758 to 0.787 (Table 2, Table S2). The Montreal Neurological Institute (MNI) coordinates of brain regions related to significant within-network connectivity are reported in Table S3.

**Table 2.** Group comparisons of triple network models using the discrimination approach*.

| Model | Bandwidth | Depression vs. HV |  |  |  | TRD vs. noTRD vs. HV |  |  |  | TRD vs. HV |  |  |  |
| --- | --- | --- | --- | --- | --- | --- | --- | --- | --- | --- | --- | --- | --- |
|  |  | Component 1 |  | Component 2 |  | Component 1 |  | Component 2 |  | Component 1 |  | Component 2 |  |
|  |  | AUROC | P-value | AUROC | P-value | AUROC | p-value | AUROC | p-value | AUROC | P-value | AUROC | P-value |
| **Maximum Discrimination Approach** | Theta | .627 | .037 | .762 | <.001 | .625 (TRD)<br>.700 (noTRD)<br>.587 (HV) | .067 (TRD)<br>.155 (noTRD)<br>.228 (HV) | .716 (TRD)<br>.763 (noTRD)<br>.670 (HV) | .009 (TRD)<br>.084 (noTRD)<br>.072 (HV) | .691 | .002 | .760 | <.001 |
|  | Alpha | .644 | .009 | .758 | <.001 | .537 (TRD)<br>.759 (noTRD)<br>.576 (HV) | .524 (TRD)<br>.039 (noTRD)<br>.212 (HV) | .649 (TRD)<br>.892 (noTRD)<br>.748 (HV) | .089 (TRD)<br>.010 (noTRD)<br>.004 (HV) | .665 | .007 | .737 | <.001 |
|  | Beta | .641 | .024 | .787 | <.001 | .654 (TRD)<br>.598 (noTRD)<br>.630 (HV) | .014 (TRD)<br>.383 (noTRD)<br>.035 (HV) | .739 (TRD)<br>.647 (noTRD)<br>.710 (HV) | <.001 (TRD)<br>.021 (noTRD)<br><.001 (HV) | .622 | .057 | .807 | <.001 |
|  | Gamma | .665 | .004 | .778 | <.001 | .602 (TRD)<br>.672 (noTRD)<br>.630 (HV) | .164 (TRD)<br>.185 (noTRD)<br>.006 (HV) | .688 (TRD)<br>.698 (noTRD)<br>.703 (HV) | .013 (TRD)<br>.118 (noTRD)<br>.006 (HV) | .697 | .003 | .758 | <.001 |

\*Triple network models consist of the default mode network (DMN), executive control network (ECN), and salience network (SN).
HV: healthy volunteer; TRD: treatment-resistant depression; noTRD: diagnosed with depression but not treatment-resistant depression; AUROC: area under the receiver operating curve

### DIABLO: Contribution of networks

To assess the contribution of each network to group differentiation, simulated ROC scores for each network were compared across different frequency bands. Model performance varied by network and bandwidth (Table S4). In the depression model, the ECN showed superior performance to the DMN and SN in the theta and beta bands in distinguishing between the depression and HV groups (ps<.05). In the TRD model, the DMN outperformed the ECN and the SN in the alpha and beta bands (ps<.05). These findings indicate that model performance depended on the specific comparison groups and frequency bands, with the ECN and DMN in the beta band showing promise for distinguishing the noTRD and TRD groups, respectively, relative to other networks.

### DIABLO: network model loadings

The DIABLO results identified key features—specifically, the most important brain connectivities—for differentiating groups. Age, sex, and race/ethnicity-adjusted MANOVA was then conducted to investigate associations between group membership and within-network connectivity. Only significant associations are reported (Table 3).

**Table 3.** Group differences among within-network connectivity scores of the triple network maximum discrimination models*

| Comparison | Bandwidths | Canonical Networks | Connectivity |  | F-value | P-value | FDR P-value | Mean: Group1 | Mean: Group2 | Diff Stats | Diff P-value | FDR Diff P-value |
| --- | --- | --- | --- | --- | --- | --- | --- | --- | --- | --- | --- | --- |
| **Depression vs. HV** | **Beta** | **ECN** | OFC | precuneus | 7.53 | .007 | .351 | 0.16 | -0.13 | 0.29 | .008 | .420 |
|  |  |  | OFC | Temp | 5.42 | .021 | .425 | 0.15 | -0.06 | 0.21 | .025 | .488 |
|  |  |  | OFC | mPFC | 4.58 | .034 | .440 | 0.16 | -0.09 | 0.25 | .039 | .488 |
| **TRD vs. HV** | **Alpha** | **SN** | SPL | FrOper | 4.57 | .034 | .068 | 0.01 | -0.2 | 0.22 | .042 | .084 |
|  | **Beta** | **DMN** | IPL | precuneus | 12.80 | <.001 | .033 | 0.23 | -0.16 | 0.39 | <.001 | .031 |
|  |  |  | IPL | PFCl | 8.32 | .005 | .315 | 0.15 | -0.15 | 0.30 | .005 | .373 |
|  |  |  | IPL | PFCd | 6.79 | .010 | .706 | 0.11 | -0.15 | 0.26 | .013 | .928 |
|  |  |  | TempPole | PHC | 6.19 | .014 | .975 | 0.12 | -0.17 | 0.29 | .015 | 1 |
|  |  | **ECN** | FPole | CingP | 6.58 | .011 | .305 | 0.14 | -0.14 | 0.28 | .012 | .335 |
|  |  |  | Temp | CingP | 4.35 | .039 | .349 | 0.21 | 0.001 | 0.21 | .047 | .421 |
|  |  |  | IPS | PFCl | 4.86 | .029 | .349 | 0.09 | -0.16 | 0.24 | .044 | .421 |
|  |  | **SN** | INS | PFCl | 7.47 | .007 | .310 | 0.16 | -0.16 | 0.32 | .009 | .395 |
|  |  |  | IFG | FrMed | 3.98 | .048 | .498 | 0.16 | -0.06 | 0.22 | .043 | .589 |
|  | **Gamma** | **SN** | Temp | INS | 4.60 | .034 | .592 | 0.18 | -0.02 | 0.20 | .028 | .591 |

\*Triple network models consist of the DMN, ECN, and SN.
HV: healthy volunteer; TRD: treatment-resistant depression; ECN: executive control network; SN: salience network; DMN: default mode network; OFC: orbitofrontal cortex; SPL: superior parietal lobule; IPL: inferior parietal lobule; TempPole: temporal pole; FPole: frontal pole; Temp: temporal cortex; IPS: intraparietal sulcus; INS: insula; IFG: inferior frontal gyrus; mPFC: medial prefrontal cortex; FrOper: frontal operculum; PFCl: lateral prefrontal cortex; PFCd: dorsal prefrontal cortex; PHC: parahippocampal cortex; CingP: posterior cingulate cortex; FrMed: frontal medial cortex.

Overall, both the depression and TRD groups exhibited hyperconnectivity in the specified connections compared to the HVs. The depression group showed increased within-network connectivity between the orbitofrontal cortex (OFC) and the precuneus, temporal cortex, and medial prefrontal cortex (mPFC) within the ECN in the beta band relative to the HV group (Fig. 3a, upper panel). As depicted in Fig. 3a (lower panel) and 3b, the TRD group demonstrated increased brain connectivity within the SN in the alpha band (between the superior parietal lobule and frontal operculum) and within the SN in the gamma band (between the temporal cortex and insula) compared to the HV group. In the beta band, the TRD group also exhibited elevated within-network connectivity in the DMN (between the IPL and the precuneus, lateral PFC (lPFC), dorsal PFC, and between the temporal pole and parahippocampal cortex), in the ECN (between the posterior cingulate cortex and frontal pole as well as the temporal cortex, and between the intraparietal sulcus and lPFC), and in the SN (between the insula and lPFC, and between the inferior frontal gyrus and frontal medial cortex) compared to the HV group. DMN within-network connectivity between the IPL and precuneus in the beta band (*F*(1,147)=12.80, *FDR-corrected p*=.033) survived multiple comparison correction. In addition, SN within-network connectivity between the superior parietal lobule and frontal operculum in the alpha band showed a trend level effect (*F*(1,148)=4.57, *FDR-corrected p*=.068).

**Figure 3.**
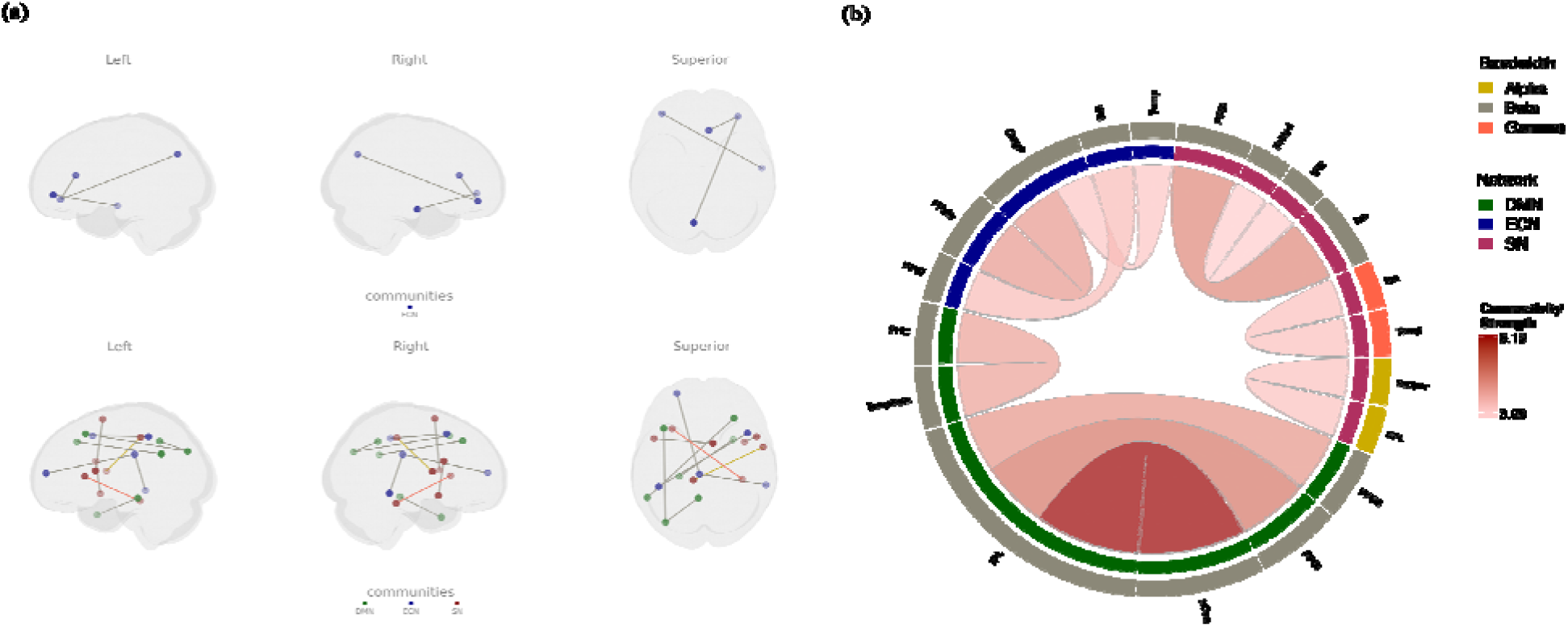
Visualization of the differences between the patient groups versus healthy volunteers (HVs). (a) Upper panel: the depression group showed hyperconnectivity in the beta band compared to the HV group; lower panel: the treatment-resistant depression (TRD) group showed hyperconnectivity in the alpha, beta, and gamma bands within the default mode network (DMN), executive control network (ECN), and salience network (SN); (b) circos plot showing differences between the TRD and HV groups across the DMN, ECN, and SN in the alpha, beta, and gamma frequencies.

### DIABLO: Loadings and treatment failures

To explore whether selected features from the TRD model were quantitatively related to the degree of treatment resistance, an ordered logistic regression analysis was conducted using the polr function in R (Tables S5, S6). Degrees of selected features were entered as predictors, while the quartile score of treatment failure frequency was used as the outcome variable (Table S6). Biological sex and age were included as covariates. Only significant findings are reported here. The results showed that connectivity between the superior parietal lobule and frontal operculum in the alpha band, and between the IPL and lPFC in the beta band, positively predicted the quantile score of treatment failure frequency (B=0.83, SE=0.32, p=.010; B=0.52, SE=0.25, p=.039, respectively). When one quartile of connectivity strength increased, participants were more likely to have severe depression, as assessed by treatment failure frequency quartile (odds=2.29 [1.22, 4.29]; odds=1.69 [1.03, 2.77], respectively). Sensitivity analyses were conducted to confirm that the associations were not influenced by MADRS score (M=32.10, SD=7.60), medication history (binomial test, 12 out of 74, p<.001), or polypharmacy (M=0.35, SD=1.11). Associations between connectivity and treatment failure frequency remained robust after adjusting for covariates (Table S7).

## Discussion

This study hypothesized that individuals with depression would exhibit increased brain connectivity within the DMN, ECN, SN, DAN, MN, and VN compared to HVs and that group differences in individuals with depression versus HVs could be predicted using a six-network model. DIABLO analyses using the so-called “triple network” model (DMN, ECN, SN) effectively distinguished the depression group from the HV group, suggesting potential markers for depression versus HVs. Notable markers included connectivity between the OFC and the precuneus, temporal cortex, and mPFC within the ECN in the beta band. Because most individuals with depression in this sample had TRD, additional analyses examined whether the triple network model could differentiate individuals with TRD from HVs. The results indicated potential within-network differences that distinguished individuals with TRD from HVs, including connectivities in the IPL, precuneus, insula, inferior frontal gyrus, frontal medial cortex, temporal gyrus, frontal/temporal pole, posterior cingulate cortex, and prefrontal cortices. Notably, hyperconnectivity between certain frontal and parietal regions was associated with severity of depressive symptoms (as assessed by the number of treatment failures). These findings highlight key brain regions and connectivity patterns, advancing our understanding of the neural mechanisms underlying depression and treatment resistance.

When the six-network model (DMN, ECN, SN, DAN, MN, VN) was compared to the triple network model (DMN, ECN, SN), the results indicated that the triple network model outperformed the six-network model in differentiating individuals with depression from HVs. While model performance alone should not be the sole criterion for model selection, recent studies similarly support the adequacy of the triple network approach for investigating depression. For instance, several reviews previously identified the DMN, ECN, and SN as central to the mechanisms underlying mental disorders, including depression [6, 49].

Interestingly, the ECN has been linked to suicidal ideation in depressed adolescents [50], and individuals with depression showed both increased connectivity within the ECN and decreased connectivity from the SN to the DMN compared to controls [51]. The same study found that connectivity between the ECN and SN correlated significantly with the severity of depressive symptoms [51]. Collectively, these findings support the sufficiency and relevance of the triple network model for elucidating the neural mechanisms underlying depression, particularly TRD. Future research should further refine this approach and investigate additional modulatory factors.

A notable aspect of our findings is the frequency-specific contribution of each canonical network in differentiating individuals with depression, especially those with TRD, from HVs. In our analyses, the ECN most effectively distinguished those with depression from HVs in the theta and beta bands, while in the TRD model, the DMN demonstrated superior discriminatory power in the beta band compared to the ECN. This suggests that each canonical network contributes uniquely to group differentiation, particularly within the theta and beta frequency bands. Moreover, previous studies identified increased theta and beta power as potential markers for depression [26–28], although comparisons between networks have not been consistently significant in systematic reviews [52, 53], and the predictive value of these frequency bands remains uncertain. Our findings advance this literature by demonstrating significant differences between networks, contributing to a more nuanced understanding of the frequency- and network-specific neural mechanisms underlying depression and, potentially, TRD.

Our results indicated increased brain connectivity within the DMN, ECN, and SN in the depression/TRD group compared to the HV group. Previous findings on this topic have been somewhat mixed. For example, one study found that individuals with remitted depression had higher within- and between-brain connectivity in these networks than those with current depression and HVs [30], while other studies reported functional disconnection within the DMN in depressed patients, particularly involving the precuneus and angular gyrus [54]. It is important to note that these studies often relied on fMRI-derived signals or averaged oscillatory coupling measures to estimate functional connectivity [54], which may differ from our MEG-based approach [26, 30]. EEG studies also found that individuals with bipolar disorder exhibited increased functional connectivity in the alpha and theta bands relative to those with depression, though spatial specificity was limited by the number of EEG channels [55]. Compared to HVs, individuals with depression have also shown increased connectivity across multiple frequency bands, especially in anterior brain regions [56]. Taken together, these studies are generally consistent with our findings of increased within-network connectivity in individuals with depression, including those with TRD. However, further research is needed to validate and clarify the role of increased within-network connectivity as a neural marker in depression.

Compared to the HV group, the TRD group exhibited hyperconnectivity in multiple brain regions. The superior parietal lobule and IPL are partially involved in sensory perception and multisensory integration [57–60], whereas the frontal operculum and lPFC have been implicated in cognitive control, emotion regulation, and stress-related functioning [61–64]. These alterations may be clinically relevant, as depression has been linked to disrupted reward-based decision making and emotion regulation [64–66], which may in turn increases vulnerability to stress [61, 63]. Thus, hyperconnectivity within the SN and DMN may reflect amplified demands on sensory and regulatory control processes in individuals with more treatment failures.

Exploratory analyses revealed widespread hyperconnectivity in the TRD group versus the noTRD group involving the OFC, IPC, posterior cingulate, insula, precuneus, parahippocampal cortex, and frontal operculum regions across the alpha and beta frequency bands. Connectivity between the superior parietal lobule and frontal operculum in the alpha band, as well as between the IPL and lPFC in the beta band, was positively associated with higher treatment failure frequency quartiles. It is worth noting that the noTRD group was relatively small compared to the TRD group (n=14 vs n=74, respectively), which may affect generalizability. These abnormalities may reflect persistent rumination [3, 67–69], cognitive rigidity [70], and impaired regulation [71]—all of which are hallmarks of treatment resistance [16, 72]. Hyperconnectivity in the frontal medial cortex, lPFC, and dorsal PFC may also exacerbate deficits in adaptive decision-making [65, 66] and emotion regulation [64], increasing vulnerability to stress [61, 63]. These results suggest that individuals with TRD may have amplified sensory perception and regulation, driven by parts of the SN and DMN. Collectively, these connectivity patterns highlight both shared and distinct neurobiological mechanisms underlying symptom severity and chronicity in depression, including TRD, and highlight the potential value of network-level biomarkers that can differentiate depression subtyping and personalized treatment.

While our results suggest that hyperconnectivity might be a feature of depression, several limitations should be acknowledged. First, the noTRD comparison group was small (n=14), potentially limiting treatment-resistance specificity. Although the TRD group (n=74) was relatively large compared with prior TRD fMRI studies (Ns ranging from 17-38), overall subgroup sizes remain modest for feature detection and predictive modeling, highlighting the need for larger studies to replicate these findings. Second, no external validation analyses were possible given the limited availability of comparable MEG datasets in TRD populations. Future collaborative efforts and open data initiatives will be essential to enable cross-dataset validation and better establish the generalizability of these findings. Lastly, group differences in age and race/ethnicity were observed, underscoring the potential confounding effects of these variables.

## Conclusion

This study demonstrated the utility of the DIABLO method for examining resting-state MEG network changes in individuals with depression compared with HVs. Collectively, the results indicate that the “triple network” model of resting brain connectivity within the DMN, ECN, and SN distinguished individuals with depression, including those with TRD, from HVs, with each network contributing uniquely across frequency bands. Increased resting brain connectivity within these networks in depression, particularly TRD, may indicate compensatory or maladaptive integration, potentially worsening deficits in decision-making and emotion regulation. The findings advance our understanding of the neural mechanisms underlying depression and treatment resistance, underscoring the value of innovative analytic approaches for future research and clinical applications.

Supplemental information is available at the *Molecular Psychiatry* website

## Data Availability

All data produced in the present study are available upon reasonable request to the authors

## Acknowledgements

This research was supported by the Intramural Research Program of the National Institutes of Health (IRP-NIMH-NIH; ZIAMH002927, conducted under clinical trials NCT00088699, NCT00369915, NCT02543983, NCT00397111, NCT03065335, NCT04821271, NCT00024635, NCT00472576, NCT00759395, and NCT02049385, and protocol 09-M-0240).

The contributions of the NIH authors are considered Works of the United States Government. The findings and conclusions presented in this paper are those of the authors and do not necessarily reflect the views of the NIH or the US Department of Health and Human Services.

The authors thank the 7SE research unit and staff for their support. Ioline Henter (NIMH) provided invaluable editorial assistance. This work utilized the computational resources of the National Institutes of Health (NIH) high-performance computing (HPC) Biowulf cluster (http://hpc.nih.gov).

## Conflict of Interest

Dr. Zarate is listed as a co-inventor on a patent for the use of ketamine in major depression and suicidal ideation; as a co-inventor on a patent for the use of (2*R*,6*R*)- hydroxynorketamine, (*S*)-dehydronorketamine, and other stereoisomeric dehydroxylated and hydroxylated metabolites of (*R,S*)-ketamine metabolites in the treatment of depression and neuropathic pain; and as a co-inventor on a patent application for the use of (2*R*,6*R*)- hydroxynorketamine and (2*S*,6*S*)-hydroxynorketamine in the treatment of depression, anxiety, anhedonia, suicidal ideation, and post-traumatic stress disorder. He has assigned his patent rights to the U.S. government but will share a percentage of any royalties that may be received by the government. All other authors have no conflict of interest to disclose, financial or otherwise.

## Notes

### Competing Interest Statement

The authors have declared no competing interest.

### Author Declarations

The IIRB of the National Institutes of Health gave ethical approval for this work

