## Supplemental Methods for "Electrophysiological Markers of Within-Network Connectivity in Major Depression"

**SUPPLEMENT**

**Supplemental Methods**

*Participants*

Participants were adults aged 18–70 years who met DSM-5 criteria for major depressive disorder (MDD) without psychotic features and had at least moderate depressive symptom severity (Montgomery-Asberg Depression Rating Scale (MADRS) score ≥ 22; Young Mania Rating Scale (YMRS) ≤ 12). Comorbid anxiety disorders were permitted. Exclusion criteria included current psychotic features or psychotic disorder; substance use disorder within the past three months; serious or unstable medical illness (e.g, hypertension, diabetes); pregnancy or breastfeeding; neurological illness or injury (e.g., seizure history, epilepsy, stroke, brain surgery, significant head injury, or structural brain lesion); medical conditions likely to affect brain morphology or physiology; clinically significant laboratory abnormalities; hearing loss relevant to imaging procedures; positive HIV test; weight >119 kg; opioid use within the past 3 months; unwillingness to discontinue structured psychotherapy; MRI/MEG contraindications (e.g., ferromagnetic implants or claustrophobia); current serious suicidal or homicidal risk; history of aggressive behavior; or current NIMH employment or NIMH employment of an immediate family member.

*DIABLO analysis*

Data Integration Analysis for Biomarker Discovery using Latent Variable Approaches for Omics Studies (DIABLO), implemented in the mixOmics package in R, was used to identify within-network neural connectivity that might differentiate between participant groups. DIABLO is a supervised multi-block framework designed for high-dimensional datasets. It identifies a limited set of correlated features that predict a specific outcome, making it particularly useful when the number of variables exceeds the number of samples. In this case, conventional models may fail to converge or yield unstable estimates. DIABLO addresses this issue by implementing sparse partial least squares discriminant analysis (sPLS-DA) within a latent variable framework, using feature selection and regularization to improve model stability. Conceptually, DIABLO extends sparse generalized canonical correlation analysis and projection to latent structures by integrating multiple data blocks while maximizing covariance across blocks and discrimination of the outcome. In each model, the input blocks consisted of scaled within-network connectivity measures from each network (default mode network (DMN), executive control network (ECN), salience network (SN), dorsal attention network (DAN), motor network (MN), and visual network (VN)), and the outcome variable encoded group membership (depression vs. healthy volunteer (HV); treatment-resistant depression (TRD) vs noTRD vs HV; TRD vs HV). This procedure results in a multivariate signature of correlated features across blocks measured in the same individual that most discriminates the outcome of interest. To examine frequency-specific neural contributions and improve interpretability, DIABLO analyses were conducted separately within each MEG frequency band (theta (4-8 Hz), alpha (9-14 Hz), beta (15-29 Hz), and gamma (30-58 Hz)).

Three parameters were tuned: the number of components, the design matrix, and the number of blocks. The optimal number of components, representing the latent dimensions that most explained group membership, was determined using the perf function. The optimal number of within connectivity features retained from each network was selected via an iterative grid search using the tune.block.splsda function [1].

To determine whether model performance was improved by emphasizing inter-network integration or class discrimination, two approaches were compared for specifying the DIABLO design matrix in the primary depression vs. HV analysis using the triple-network model (DMN, ECN, and SN). First, correlational structure was estimated among networks using the sparse approach based on sPLS. Correlations from the first component of each sparse model were used to inform the DIABLO design matrix, with values ranging from 0 to 1 that represented the degree of correlation to be modeled between data blocks. DIABLO model based on the design matrix was fit and evaluated using the area under the receiver operating curve (AUROC) with centroid-based, five-fold, 100 repeated cross-validation, implemented with the auroc function in the DIABLO package. A maximum discrimination model was then fit by fixing off-diagonal elements of the design matrix to 0.1, thereby placing less emphasis on inter-block correlation and more emphasis on class discrimination [1-3]. The discriminative contribution of each selected connectivity feature was then examined. This comparison allowed us to assess whether group discrimination was better supported by an integrative model reflecting empirical relationships among networks or by a more discrimination-oriented model.

Because the number of input blocks can influence both computational burden and classification performance, a triple network model (DMN, ECN, and SN) and a six network model (DMN, ECN, SN, DAN, MN, and VN) were compared across group differences (depression vs HV; TRD vs noTRD vs HV; TRD vs HV) to determine whether the canonical depression-related networks were sufficient for group discrimination or whether additional networks improved predictive performance. Model performance was evaluated using AUROC with the same settings. Because individuals with TRD comprised most of the depression group, differences between the depression and HV groups were likely driven largely by TRD. A simulation-based bootstrap approach was used to compare predictive accuracy across networks within a given frequency band. Specifically, binary classification outcomes were simulated for each network (DMN, ECN, and SN) based on the observed AUROC values, assuming binomial sampling of predictions. For each pairwise comparison, we generated simulated receiver operating curve (ROC) and tested whether the difference in simulated ROC was statistically significant using the bootstrap method implemented in the roc.test function from the pROC package in R [4], with 10,000 bootstrap resamples.

With reasonable performance, the significantly contributing brain connectivity scores in each canonical network for the first and second components, which most effectively differentiated group membership, were exported and used for follow-up analyses. Due to the nature of latent-based DIABLO analysis, outcomes were estimated for two latent components (linear combinations of connectivity features designed to maximize group discrimination). Component 1 captured the greatest shared variance and group discrimination across groups, while Component 2 explained additional independent variance and discriminatory information. Only AUROC scores for Component 2 are reported, as it explained independent variance and enhanced discriminative accuracy beyond what was captured by Component 1 alone.

**Supplementary References**

1.	Lê Cao K-A, Welham ZM. *Multivariate data integration using R: methods and applications with the mixOmics package*. Chapman and Hall/CRC: New York, 2021.

2.	Burton‐Pimentel KJ, Pimentel G, Hughes M, Michielsen CC, Fatima A, Vionnet N *et al.* Discriminating dietary responses by combining transcriptomics and metabolomics data in nutrition intervention studies. *Mol Nutr Food Res* 2021; **65**(4)**:** 2000647.

3.	Rohart F, Gautier B, Singh A, Lê Cao K-A. mixOmics: An R package for ‘omics feature selection and multiple data integration. *PLoS Comput Biol* 2017; **13**(11)**:** e1005752.

4.	Robin X, Turck N, Hainard A, Tiberti N, Lisacek F, Sanchez J-C *et al.* pROC: an open-source package for R and S+ to analyze and compare ROC curves. *BMC Bioinformatics* 2011; **12:** 77.

**Supplementary Table S1. Six network model* results**

| **Model** | **Band-**  **width** | **Depression vs. HV** | | | | **TRD vs. noTRD vs. HV** | | | | **TRD vs. HV** | | | |
| --- | --- | --- | --- | --- | --- | --- | --- | --- | --- | --- | --- | --- | --- |
|  |  | **Component 1** | | **Component 2** | | **Component 1** | | **Component 2** | | **Component 1** | | **Component 2** | |
|  |  | **AUC** | **p-value** | **AUC** | **p-value** | **AUC** | **p-value** | **AUC** | **p-value** | **AUC** | **p-value** | **AUC** | **p-**  **value** |
| **Maximum Discrimination Approach** | Theta | .625 | .036 | .761 | <.001 | .604 (TRD) .685 (noTRD) .566 (HV) | .100 (TRD) .128 (noTRD) .307 (HV) | .722 (TRD) .774 (noTRD) .680 (HV) | .023 (TRD) .054 (noTRD) .041 (HV) | .648 | .041 | .797 | <.001 |
|  | Alpha | .622 | .033 | .754 | <.001 | .526 (TRD) .762 (noTRD) .570 (HV) | .612 (TRD) .027 (noTRD) .234 (HV) | .581 (TRD) .876 (noTRD) .668 (HV) | .238 (TRD) .006 (noTRD) .019 (HV) | .634 | .069 | .783 | <.001 |
|  | Beta | .600 | .118 | .762 | <.001 | .579 (TRD) .603 (noTRD) .559 (HV) | .206 (TRD) .359 (noTRD) .316 (HV) | .698 (TRD) .689 (noTRD) .665 (HV) | .004 (TRD) .150 (noTRD) .036 (HV) | .609 | .077 | .770 | <.001 |
|  | Gamma | .645 | .021 | .747 | <.001 | .551 (TRD) .647 (noTRD) .580 (HV) | .393 (TRD) .180 (noTRD) .202 (HV) | .676 (TRD) .666 (noTRD) .702 (HV) | .006 (TRD) .135 (noTRD) .001 (HV) | .602 | .128 | .728 | .001 |

*The six network model consists of the default mode network (DMN), executive control network (ECN), salience network (SN), dorsal attention network (DAN), motor network (MN), and visual network (VN).

HV: healthy volunteer; TRD: treatment-resistant depression; noTRD: diagnosed with depression but not treatment-resistant depression; AUC: area under the curve

**Supplementary Table S2. Two approaches for comparing the depression and healthy volunteer groups using triple network models***

| **Model** | **Bandwidth** | **Depression vs. HV** | | | |
| --- | --- | --- | --- | --- | --- |
|  |  | **Component 1** | | **Component 2** | |
|  |  | **AUC** | **p-value** | **AUC** | **p-value** |
| **Sparse Approach** | Theta | .552 | .364 | .625 | .083 |
|  | Alpha | .595 | .114 | .646 | .026 |
|  | Beta | .556 | .320 | .611 | .081 |
|  | Gamma | .578 | .168 | .637 | .019 |
| **Maximum**  **Discrimination Approach** | Theta | .627 | .037 | .762 | <.001 |
|  | Alpha | .644 | .009 | .758 | <.001 |
|  | Beta | .641 | .024 | .787 | <.001 |
|  | Gamma | .665 | .004 | .778 | <.001 |

HV: healthy volunteer; AUC: area under the curve

*The triple network models consist of the default mode network (DMN), executive control network (ECN), and salience network (SN).

**Supplementary Table S3.** **MNI coordinates for connectivity seeds derived from group-differential vertex centroids**

| **Comparison** | **Network** | **Bandwidth** | **Hemisphere** | **Name** | **MNI coordinates** | | |
| --- | --- | --- | --- | --- | --- | --- | --- |
|  |  |  |  |  | **X** | **Y** | **Z** |
| **Depression vs. HV** | **ECN** | **Beta** | Right | Temporal cortex | 63 | -41 | -12 |
|  |  |  | Left | Orbitofrontal cortex | -42 | 49 | -6 |
|  |  |  | Right | Orbitofrontal cortex | 36 | 46 | -13 |
|  |  |  | Right | Temporal cortex | 61 | -13 | -21 |
|  |  |  | Right | Medial prefrontal cortex | 6 | 29 | 15 |
|  |  |  | Left | Precuneus | -9 | -73 | 38 |
| **TRD vs. noTRD** | **ECN** | **Alpha** | Right | Orbitofrontal cortex | 36 | 46 | -13 |
|  |  |  | Right | Lateral prefrontal cortex | 41 | 33 | 37 |
|  |  | **Beta** | Right | Lateral prefrontal cortex | 41 | 33 | 37 |
|  |  |  | Left | Medial prefrontal cortex | -6 | 30 | 25 |
| **TRD vs. HV** | **DMN** | **Beta** | Right | Dorsal prefrontal cortex | 29 | 30 | 42 |
|  |  |  | Right | Temporal pole | 30 | 9 | -38 |
|  |  |  | Left | Inferior parietal lobule | -57 | -54 | 28 |
|  |  |  | Left | Lateral prefrontal cortex | -40 | 19 | 49 |
|  |  |  | Left | Inferior parietal lobule | -39 | -80 | 31 |
|  |  |  | Left | Precuneus | -6 | -54 | 42 |
|  |  |  | Left | Parahippocampal cortex | -26 | -32 | -18 |
|  | **ECN** | **Beta** | Left | Intraparietal sulcus | -45 | -42 | 46 |
|  |  |  | Right | Temporal cortex | 63 | -41 | -12 |
|  |  |  | Right | Lateral prefrontal cortex | 42 | 14 | 49 |
|  |  |  | Left | Posterior cingulate cortex | -5 | -29 | 28 |
|  |  |  | Left | Frontal pole | -28 | 58 | 8 |
|  | **SN** | **Alpha** | Left | Superior parietal lobule | -11 | -35 | 46 |
|  |  |  | Right | Frontal operculum | 59 | 0 | 10 |
|  |  | **Beta** | Right | Medial frontal cortex | 8 | 3 | 66 |
|  |  |  | Right | Insula | 41 | 6 | -15 |
|  |  |  | Right | Lateral prefrontal cortex | 52 | 11 | 21 |
|  |  |  | Left | Inferior frontal gyrus | -51 | 9 | 11 |
|  |  | **Gamma** | Left | Insula | -33 | 20 | 5 |
|  |  |  | Right | Temporal cortex | 39 | -35 | -23 |

Depression including TRD and noTRD. HV: healthy volunteer; TRD: treatment-resistant depression; noTRD: diagnosed with depression but not treatment-resistant depression; MNI: Montreal Neurological Institute; DMN: default mode network; ECN: executive control network; SN: salience network

**Supplementary Table S4. Contribution of each network to the maximum difference using triple network models*** **in different bandwidths**

| **Model** | **Band-**  **widths** | **Default Mode Network (DMN)** | | | | | **Executive Control Network (ECN)** | | | | | **Salience Network (SN)** | | | | **Comparison** | |
| --- | --- | --- | --- | --- | --- | --- | --- | --- | --- | --- | --- | --- | --- | --- | --- | --- | --- |
|  |  | **Component 1** | | | **Component 2** | | **Component 1** | | | **Component 2** | | **Component 1** | | **Component 2** | | **Component 2** | |
|  |  | **AUC** | **P-value** | **AUC** | | **P-value** | **AUC** | **P-value** | **AUC** | | **P-value** | **AUC** | **P-value** | **AUC** | **P-value** | **Diff** | **P-value** |
| **Depression vs. HV** | Theta | .626 | .005 | .678 | | <.001 | .647 | <.001 | **.856** | | <.001 | .592 | .039 | .695 | <.001 | -6.16(D vs. E) -2.27 (D vs. S) 5.52 (E vs. S) | <.001 .023 <.001 |
|  | Alpha | .641 | .002 | .738 | | <.001 | .637 | .002 | .700 | | <.001 | .630 | .004 | **.767** | <.001 | 2.49 (D vs. E) -2.49 (D vs. S) -3.60 (E vs. S) | .013 .013 <.001 |
|  | Beta | .623 | .006 | .727 | | <.001 | .653 | .001 | **.826** | | <.001 | .627 | .005 | .692 | <.001 | -4.20 (D vs. E) 2.48 (D vs. S) 5.03 (E vs. S) | <.001 .013 <.001 |
|  | Gamma | .619 | .007 | .697 | | <.001 | .633 | .003 | **.777** | | <.001 | .662 | <.001 | **.775** | <.001 | -3.91 (D vs. E) -3.75 (D vs. S) 1.00 (E vs. S) | <.001 <.001 .317 |
| **TRD vs. HV** | Theta | .738 | <.001 | **.767** | | <.001 | .643 | .002 | .692 | | <.001 | .668 | <.001 | **.753** | <.001 | 3.45 (D vs. E) 1.01 (D vs. S) -3.26 (E vs. S) | .001 .314 .001 |
|  | Alpha | .696 | <.001 | **.759** | | <.001 | .659 | .001 | .696 | | <.001 | .629 | .006 | .710 | <.001 | 3.11 (D vs. E) 2.70 (D vs. S) -3.11 (E vs. S) | .002 .007 .002 |
|  | Beta | .581 | .085 | **.849** | | <.001 | .551 | .277 | .707 | | <.001 | .640 | .003 | .773 | <.001 | 5.03 (D vs. E) 3.45 (D vs. S) -3.43 (E vs. S) | <.001 <.001 .001 |
|  | Gamma | .694 | .003 | .694 | | <.001 | .730 | <.001 | **.774** | | <.001 | .673 | <.001 | .699 | <.001 | -3.61 (D vs. E) -1.43 (D vs. S) 3.61 (E vs. S) | <.001 .153 <.001 |

*Triple network models consist of the default mode network (DMN), executive control network (ECN), and salience network (SN)

TRD: treatment-resistant depression; HV: healthy volunteer; AUC: area under the curve

**Supplementary Table S5. Loadings and treatment failure frequencies**

| **Model** | | **TRD vs. HV** | | | | | | | | | | |
| --- | --- | --- | --- | --- | --- | --- | --- | --- | --- | --- | --- | --- |
| **Bandwidths** | | **Alpha** | **Beta** | | | | | | | | | **Gamma** |
| **Canonical Networks** | | **SN** | **DMN** | | | | **ECN** | | | **SN** | | **SN** |
| **Connectivity** | | **SPL** | **IPL** | **IPL** | **IPL** | **TempPole** | **FPole** | **Temp** | **IPS** | **Ins** | **IFG** | **Temp** |
|  |  | **FrOper** | **precuneus** | **PFCl** | **PFCd** | **PHC** | **CingP** | **CingP** | **PFCl** | **PFCl** | **FrMed** | **Ins** |
| **AIC** | | 214.58 | 219.31 | 217.09 | 220.35 | 220.50 | 221.47 | 220.88 | 221.51 | 220.12 | 220.54 | 220.16 |
| **Connectivity Scores** | **Beta** | 0.83 | 0.40 | 0.52 | 0.34 | 0.25 | 0.06 | 0.20 | 0.04 | 0.31 | 0.27 | 0.39 |
|  | **SE** | 0.32 | 0.27 | 0.25 | 0.32 | 0.25 | 0.25 | 0.25 | 0.25 | 0.26 | 0.27 | 0.33 |
|  | **p-value** | **.010** | .139 | **.039** | .278 | .312 | .798 | .419 | .864 | .236 | .319 | .242 |
|  | **Odds ratio** | 2.29 | 1.50 | 1.69 | 1.41 | 1.28 | 1.07 | 1.22 | 1.04 | 1.37 | 1.30 | 1.47 |
|  | **Odds Low** | 1.22 | 0.88 | 1.03 | 0.76 | 0.79 | 0.65 | 1.22 | 0.64 | 0.82 | 0.77 | 0.77 |
|  | **Odds High** | 4.29 | 2.56 | 2.77 | 2.62 | 2.09 | 1.75 | 1.99 | 1.70 | 2.29 | 2.20 | 2.82 |
| **Biological Sex** | **Beta** | 0.74 | 0.86 | 0.76 | 0.74 | 0.71 | 0.69 | 0.72 | 0.67 | 0.80 | 0.73 | -0.004 |
|  | **SE** | 0.43 | 0.44 | 0.42 | 0.43 | 0.42 | 0.43 | 0.42 | 0.42 | 0.44 | 0.43 | 0.02 |
|  | **p-value** | .084 | .050 | .075 | .083 | .091 | .108 | .091 | .110 | .066 | .086 | .801 |
|  | **Odds ratio** | 2.09 | 2.37 | 2.13 | 2.09 | 2.04 | 1.99 | 2.05 | 1.95 | 2.24 | 2.08 | 1.00 |
|  | **Odds Low** | 0.90 | 1.00 | 0.93 | 0.91 | 0.89 | 0.86 | 0.89 | 0.86 | 0.95 | 0.90 | 0.96 |
|  | **Odds High** | 4.82 | 5.64 | 4.90 | 4.81 | 4.66 | 4.58 | 4.71 | 4.45 | 5.28 | 4.81 | 1.03 |
| **Age** | **Beta** | -0.01 | -0.01 | -0.01 | -0.01 | -0.01 | -0.004 | -0.003 | -0.004 | -0.007 | -0.01 | 0.72 |
|  | **SE** | 0.02 | 0.02 | 0.02 | 0.02 | 0.02 | 0.02 | 0.02 | 0.02 | 0.02 | 0.02 | 0.42 |
|  | **p-value** | .646 | .657 | .521 | .691 | .715 | .812 | .850 | .798 | .684 | .632 | .089 |
|  | **Odds ratio** | 0.99 | 0.99 | 0.99 | 0.99 | 0.99 | 1.00 | 1.00 | 1.00 | 0.99 | 0.99 | 2,05 |
|  | **Odds Low** | 0.96 | 0.96 | 0.96 | 0.96 | 0.96 | 0.96 | 0.97 | 0.96 | 0.96 | 0.96 | 0.89 |
|  | **Odds High** | 1.03 | 1.03 | 1.02 | 1.03 | 1.03 | 1.03 | 1.03 | 1.03 | 1.03 | 1.03 | 4.71 |

TRD: treatment-resistant depression; HV: healthy volunteer; DMN: default mode network; ECN: executive control network; SN: salience network; AIC: Akaike information criteria; SPL: superior parietal lobule; IPL: inferior parietal lobule; TempPole: temporal pole; FPole: frontal pole; Temp: temporal cortex; IPS: intraparietal sulcus; INS: insula; IFG: inferior frontal gyrus; FrOper: frontal operculum; PFCl: lateral prefrontal cortex; PFCd: dorsal prefrontal cortex; PHC: parahippocampal cortex; CingP: posterior cingulate cortex; FrMed: frontal medial cortex.

**Supplementary Table S6. Descriptive statistics of the treatment failure quartile scores**

| **Category** | **Sample Size** | **Min.** | **Max.** |
| --- | --- | --- | --- |
| **Q1** | 20 | 2 | 3 |
| **Q2** | 20 | 3 | 4 |
| **Q3** | 19 | 4 | 6 |
| **Q4** | 19 | 6 | 12 |

**Supplementary Table S7.** **Sensitivity analyses for the association between within-network connectivity and quantile scores of treatment failure frequency, adjusting for covariates**

|  |  |  | **Treatment Failure**  **Quantile Score** | **Age** | **Biological Sex** | **Covariate** |
| --- | --- | --- | --- | --- | --- | --- |
| **Superior parietal lobule & Frontal operculum** | **MADRS** | **Eff Size** | 0.98 | -0.004 | 0.71 | 0.07 |
|  |  | **SE** | 0.35 | 0.02 | 0.45 | 0.03 |
|  |  | **p-value** | .005 | .797 | .112 | .021 |
|  |  | **Eff Size**  **95%CI** | 0.01, 0.29 | -0.04, 0.80 | -0.17, 0.11 | 0.01, 0.14 |
|  |  | **Odds** | 1.66 | 0.03 | 1.58 | 0.14 |
|  |  | **Odds**  **95%CI** | 1.34, 2.65 | 0.97, 1.00 | 0.85, 2.03 | 1.01, 1.08 |
|  | **Medication**  **History** | **Eff Size** | 0.87 | -0.002 | 0.74 | -0.93 |
|  |  | **SE** | 0.32 | 0.02 | 0.43 | 0.55 |
|  |  | **p-value** | .008 | .887 | .086 | .092 |
|  |  | **Eff Size**  **95%CI** | 0.01, 0.23 | -0.04, 0.89 | -0.10, 0.09 | -2.03, 0.09 |
|  |  | **Odds** | 1.50 | 0.03 | 1.58 | 0.15 |
|  |  | **Odds**  **95%CI** | 1.50, 2.38 | 0.03, 1.00 | 1.58, 2.09 | 0.15, 0.39 |
|  | **Poly-**  **pharmacy** | **Eff Size** | 0.85 | -0.004 | 0.76 | -0.16 |
|  |  | **SE** | 0.32 | 0.02 | 0.43 | 0.17 |
|  |  | **p-value** | .009 | .832 | .076 | .328 |
|  |  | **Eff Size**  **95%CI** | 0.01, 0.22 | -0.04, 0.83 | -0.09, 0.08 | -0.50, 0.33 |
|  |  | **Odds** | 1.48 | 0.03 | 1.60 | 0.17 |
|  |  | **Odds**  **95%CI** | 1.24, 2.33 | 0.96, 1.00 | 0.92, 2.14 | 0.61, 0.85 |
| **Inferior parietal lobule & lateral Prefrontal cortex** | **MADRS** | **Eff Size** | 0.58 | -0.01 | 0.76 | 0.06 |
|  |  | **SE** | 0.26 | 0.02 | 0.45 | 0.03 |
|  |  | **p-value** | .028 | .600 | .089 | .067 |
|  |  | **Eff Size**  **95%CI** | 0.06, 1.10 | -0.04, 0.03 | -0.12, 1.63 | -0.004, 0.12 |
|  |  | **Odds** | 1.78 | 0.99 | 2.14 | 1.06 |
|  |  | **Odds**  **95%CI** | 1.06, 2.99 | 0.96, 1.03 | 0.89, 5.12 | 1.00, 1.13 |
|  | **Medication**  **History** | **Eff Size** | 0.52 | -0.01 | 0.74 | -0.79 |
|  |  | **SE** | 0.25 | 0.02 | 0.43 | 0.55 |
|  |  | **p-value** | .043 | .693 | .082 | .148 |
|  |  | **Eff Size**  **95%CI** | 0.02, 1.01 | -0.04, 0.03 | -0.09, 1.58 | -1.86, 0.28 |
|  |  | **Odds** | 1.67 | 0.99 | 2.10 | 0.45 |
|  |  | **Odds**  **95%CI** | 1.02, 2.74 | 0.96, 1.03 | 0.91, 4.85 | 0.06, 1.32 |
|  | **Poly-**  **pharmacy** | **Eff Size** | 0.53 | -0.01 | 0.77 | -0.15 |
|  |  | **SE** | 0.25 | 0.02 | 0.43 | 0.17 |
|  |  | **p-value** | .035 | .670 | .070 | .355 |
|  |  | **Eff Size**  **95%CI** | 0.04, 1.03 | -0.04, 0.03 | -0.06, 1.61 | -0.48, 0.17 |
|  |  | **Odds** | 1.70 | 0.99 | 2.17 | 0.86 |
|  |  | **Odds**  **95%CI** | 1.04, 2.80 | 0.96, 1.03 | 0.94, 5.01 | 0.62, 1.19 |

MADRS: Montgomery-Asberg Depression Rating Scale; CI: confidence interval; SE: standard error
